# Rapid Rise of S-Gene Target Failure and the UK variant B.1.1.7 among COVID-19 isolates in the Greater Toronto Area, Canada

**DOI:** 10.1101/2021.02.09.21251225

**Authors:** Kevin A. Brown, Jonathan Gubbay, Jessica Hopkins, Samir Patel, Sarah A. Buchan, Nick Daneman, Lee Goneau

## Abstract

A novel variant of SARS-CoV-2, B.1.1.7, originally discovered in the United Kingdom (UK), is rapidly overtaking other strains around the globe. In certain assays, absence of detection of the S-gene target, also known as S-gene target failure (SGTF) can be a sensitive surrogate of B.1.1.7. We analyzed daily counts of SGTF among samples from Dynacare Laboratory Ontario (which draws samples from the Greater Toronto Area) and resulted between December 16, 2020 and February 3, 2021. We identified 11,485 positive COVID-19 tests, of which 448 had SGTF (3.9%). The estimated prevalence of SGTF rose from 2.0% on December 16 to 15.2% on February 3 (1.8-fold weekly increase, 95%CI: 1.5, 2.2). The estimated reproduction number for SGTF cases was 1.17 (95%CI: 0.94 to 1.46), while the reproduction number for non-SGTF cases was 0.82 (95%CI: 0.65 to 1.01); the relative reproduction number was 1.44 (95%CI: 1.03, 1.99). 59 samples were sent for confirmatory testing, of which 53 (90%) were identified as B.1.1.7 using whole genome sequencing or found to have the N501Y mutation. In order to pre-emptively plan and implement public health measures to control COVID-19 now and through the spring, accurate and up-to-date early warning systems for new variants, including B.1.1.7, are essential across North America and the globe.

## Background

A novel variant of SARS-CoV-2, B.1.1.7, originally discovered in the United Kingdom (UK), is rapidly overtaking other strains around the globe,^1^ due to a substantial transmission advantage; the B.1.1.7 variant is estimated to be 40-80% more transmissible than wild-type strains.^2^ Preliminary data also suggest a 1.35-fold higher probability of mortality associated with the B.1.1.7 variant.^3^

In certain assays, absence of detection of the S-gene target, also known as S-gene target failure (SGTF) can be a sensitive surrogate of B.1.1.7. However, prior to B.1.1.7 introduction into a jurisdiction, other factors can erroneously trigger SGTF, leading to low specificity. These include samples with low overall viral burden (Cycle threshold [Ct] values > 30), or other, non-B.1.1.7 variants possessing polymorphisms within the primer or probe binding of S gene (e.g., 69-70 deletion)^4^.Prior to strain dominance, a consistent, 10-fold monthly increase in B.1.1.7 prevalence was identified in regions of England.^2^ Rapid increases in SGTF prevalence is currently thought to be a strong signal of strain replacement with B.1.1.7, after validation with representative genome sequencing of local specimens.

To track and quantify the potential spread of B.1.1.7 in Ontario, we examined the incidence of SGTF, and conducted confirmatory testing, in the December 2020 to February 2021 period.

## Methods

We analyzed daily counts SGTF among samples tested using the TaqPath COVID-19 Combo Kit from Dynacare Laboratory Ontario (which draws samples from the Greater Toronto Area, in particular: Markham, Brampton, Maple, and Etobicoke) and resulted between December 15, 2020 and February 3, 2021. SGTF was defined as non-detection of the S-gene target among samples that were positive (Ct < 37) for both N gene and *orf1ab* targets. Based on these daily counts, we fit regression models to examine rate of change in the proportion of SGTF. We also examined the reproduction number of SGTF, non-SGTF, and a subset of non-outbreak associated SGTF samples, using the R epinow2 package,^5^ and assuming a generation interval of 5.2 days.^6^ Aliquots of SGTF samples that possessed N gene or *orf1ab* target Ct values ≤ 30 were forwarded to Public Health Ontario for confirmatory testing.

## Results

Between December 16, 2020 and February 3, 2021, we identified 11,485 positive COVID-19 tests, of which 448 had SGTF (3.9%). Baseline SGTF prevalence was 2.0% on December 16^th^, and the prevalence of SGTF began to increase in early January 2021 (**Figure 1**). In the 3-week period since January 13, the rate of increase in SGTF prevalence was estimated to be 1.8-fold per week (incidence ratio = 1.8, 95%CI: 1.5 to 2.2, odds ratio = 1.9, 95%CI: 1.6 to 2.2). The estimated prevalence of SGTF on February 3^rd^ was 15.2%; the projected date of 50% SGTF prevalence, based on the logistic trend, was February 20^th^.

**Figure 1.**
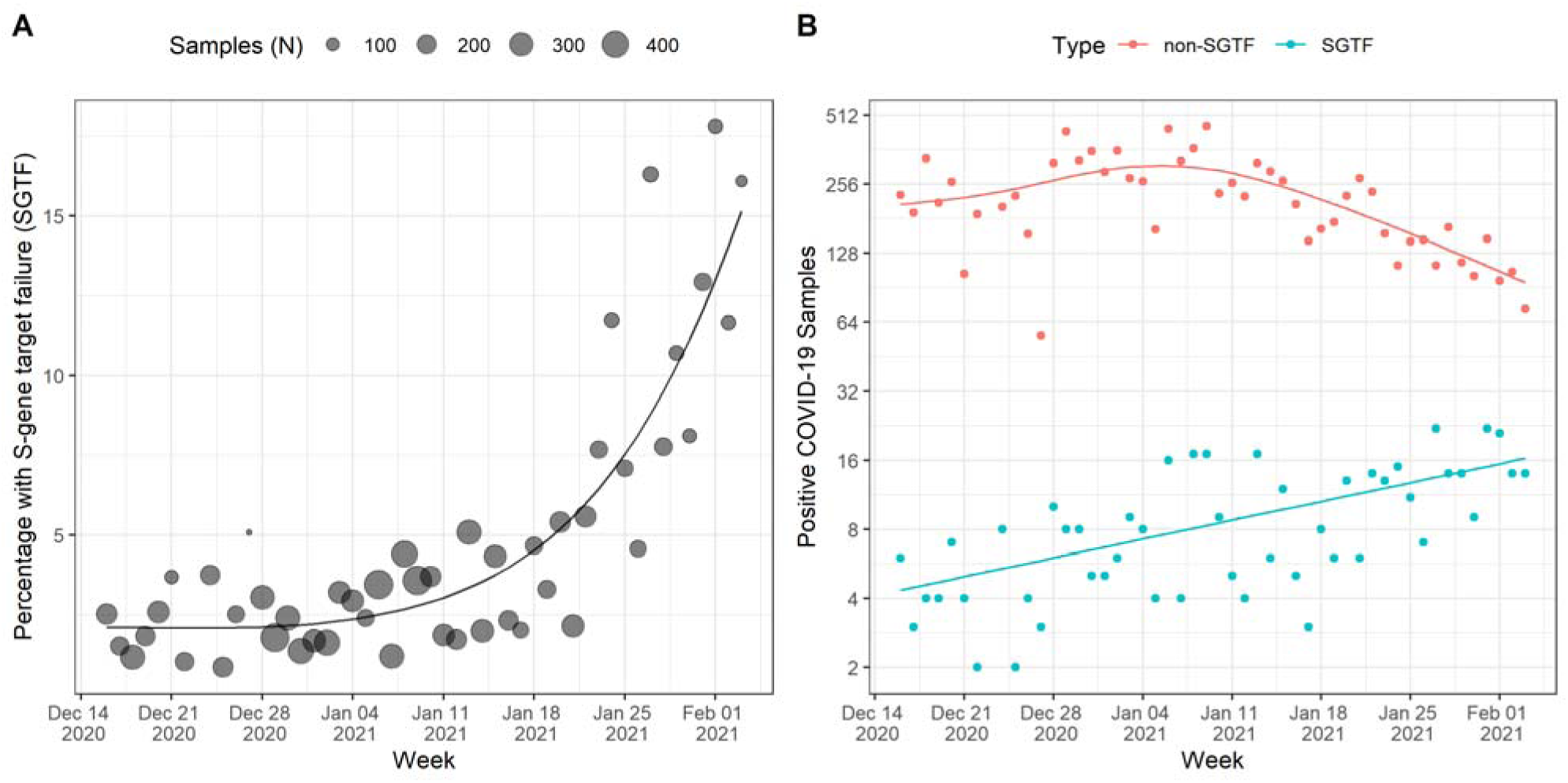
**Panel A**. Percentage of Samples with S-gene target failure (SGTF) since December 16, 2020. Over the most recent 3-weeks, a 1.8-fold increase per week was estimated. Estimated date of 50% prevalence was February 20^th^. **Panel B**. Daily Counts of SGTF versus non-SGTF Samples. Current estimated reproduction number for SGTF (R_e_=1.17) and non-SGTF (R_e_=0.82) samples are divergent (relative R_e_ = 1.44).

The estimated reproduction number for SGTF cases was 1.17 (95%CI: 0.94 to 1.46), while the reproduction number for non-SGTF cases was 0.82 (95%CI: 0.65 to 1.01); the relative reproduction number was 1.44 (95%CI: 1.03, 1.99). We also restricted our analyses to SGTF samples that were not associated with identified outbreaks. In this sensitivity analysis, we measured the reproduction number to be 1.21 (95%CI: 0.96 to 1.52).

59 samples received by Public Health Ontario had valid findings. In late December 2020, 2 of 6 SGTF samples (33%) were confirmed as B.1.1.7 using whole genome sequencing (WGS). In January 2021, 51 of 53 SGTF samples (96%) were confirmed as B.1.1.7 using WGS or found to have the N501Y mutation.^7^

## Discussion

We found an SGTF prevalence of 15% on February 3^rd^, in the Greater Toronto Area. Consistent with other jurisdictions, we measured a 10-fold monthly increase (1.8-fold weekly) in SGTF prevalence and a relative reproduction number of 1.4.^2^ Our data are consistent with a recent Ontario-wide point prevalence survey that showed 5.5% prevalence on January 20^th^.^7^ Despite overall declines in COVID-19 incidence, we found evidence that B.1.1.7 is currently expanding in the Greater Toronto Area, and will likely overtake non-SGTF variants before the end of February.

This study has a number of limitations. These data represent a sample from a region which has experienced high rates of COVID-19, and includes outbreak-related cases, and as such is not representative of the entire province. Nevertheless, the exponential increase in case counts is cause for concern. Further, our approach to SGTF detection used all positive samples, including results with high Ct values where SGTF could be incidental. As such, our approach was likely not as specific a marker of B.1.1.7 as samples with lower Ct values (e.g., Ct values <30).

In order to pre-emptively plan and implement public health measures to control COVID-19 through the spring, accurate and up-to-date early warning systems for new variants, including B.1.1.7, are essential across North America and the globe.

## Data Availability

All primary analyses in the manuscript are based on the case counts presented in Figure 1, and can be retrieved directly from the figure.

